# Methods for Factors Affecting Behavioral Treatment in Veterans with Headache Attributable to Mild to Moderate Traumatic Brain Injury

**DOI:** 10.1101/2024.07.18.24310642

**Authors:** Paul S. Nabity, Timothy T. Houle, Dana P. Turner, Stacey Young-McCaughan, Cindy A. McGeary, Mary Jo Pugh, Donald D. McGeary

## Abstract

Over 400,000 U.S. military personnel have been diagnosed with a mild or moderate traumatic brain injury (TBI) since the year 2000. Posttraumatic headache (PTH) is one of the most common and bothersome sequela after a mild or moderate head injury. Persistent posttraumatic headache are headaches due to the head injury lasting longer than 3 months. About 40% of military personnel who develop PTH after a TBI have persistent PTH and about 20% have PTH lasting longer than a year after the original injury. Persistent PTH has a negative impact on daily activities, including work and social functioning.

There are no available guidelines for treating posttraumatic headache, and there is extraordinary variability in treatment practices as a result. The present study aims to identify predictive factors that account for heterogeneity in response to behavioral intervention for posttraumatic headache attributable to mild to moderate TBI. We intend to create a predictive model of PTH through the adoption of the Predictive Approaches to Treatment effect Heterogeneity Statement (PATH Statement). This rigorous, guided approach will be used to develop and validate a predictive model of psychosocial factors related to PTH treatment outcomes, thereby improving individualized treatment of PTH. This protocol provides an overview of the research design and methods for this study.

## 1. Introduction

Over 450,000 U.S. military personnel have been diagnosed with a mild or moderate traumatic brain injury (TBI) since the year 2000 [1]. Posttraumatic headache (PTH) is one of the most common and bothersome sequela after a mild or moderate head injury [2]. Persistent posttraumatic headache are headaches due to the head injury lasting longer than 3 months. About 50% of military personnel who develop PTH after a TBI have persistent PTH and about 30% have PTH lasting longer than a year after the original injury [3]. Persistent PTH has a negative impact on daily activities, including work and social functioning.

There are no available guidelines for treating posttraumatic headache, and there is extraordinary variability in treatment practices as a result. A global survey of physicians treating PTH found that the three most utilized interventions included reassurance, education about lifestyle factors affecting headache, and prescription of headache preventative medication [5]. Although all providers surveyed agreed on the potential benefits of non-pharmacological interventions for PTH (including cognitive and behavioral therapies for mood and biofeedback-assisted behavioral therapy for headache), only 32% recommended non-pharmacological interventions because of a lack of guidance or resources to implement them. Non-pharmacological interventions are particularly important for military veterans with PTH because military PTH is highly comorbid with posttraumatic stress disorder (PTSD) [6], which exacerbates PTH disability [7].

PTSD causes pain to be more persistent and less responsive to frontline treatments including pharmacological agents [8, 9, 10]. Medication use as a first-line strategy to address pain in military service members and veterans with comorbid pain and PTSD may elevate risk for suicide [11]. Comorbid pain and PTSD is also associated with overuse of opioid medication [12]. The extant research on non-pharmacological intervention for PTH is methodologically weak, so well-designed studies are needed [13]. Our research group completed the first large study of nonpharmacological treatment of military PTH that found significant improvement in headache disability with a behavioral headache treatment [14]. Due to heterogeneity of characteristics in veterans and service members, some individual may respond better to behavioral treatment than others.

The present study aims to identify predictive factors that account for heterogeneity in response to behavioral intervention for posttraumatic headache attributable to mild to moderate TBI. We intend to create a predictive model of PTH through the adoption of the Predictive Approaches to Treatment effect Heterogeneity Statement (PATH Statement) [15]. This rigorous, guided approach will be used to develop and validate a predictive model of psychosocial factors related to PTH treatment outcomes, thereby improving individualized treatment of PTH. This project will specify, estimate, update, and partially validate a prediction model for the heterogeneity in response to behavioral intervention for PTH attributable to mild traumatic brain injury (mTBI). This on-line protocol provides an overview of the research design and methods for this study.

### 2.1 Objective: Phase 1

Our research group completed the first large study of nonpharmacological treatment of military PTH that found significant improvement in headache disability with a behavioral headache treatment [14]. We previously specified basic prediction models assessing the contribution of baseline PTSD and baseline headache disability using the headache impact test-6-items (HIT-6 score) to determine if a simple prediction model could identify heterogeneity in nonpharmacological treatment response. We examined outcomes from our clinical trial using these simple models and were able to detect notable heterogeneity. The objective of this secondary data analysis is to use the data collected to identify predictive factors that account for heterogeneity in response to behavioral intervention for posttraumatic headache attributable to mTBI.

### 2.2 Data Sources: Phase 1

Candidate variables for the model specification and initial estimation will be based on a prior randomized controlled trial. The data is stored in a local repository and will be released without identifiers to the study team under a Repository Recipient Investigator Agreement.

### 2.3 Study Population: Phase 1

Model development will proceed from data collected as part of a completed randomized controlled trial (NCT02419131). The eligibility criteria for the analysis are the same as for this individual trial, with no additional exclusion criteria. The methods for this completed trial are explained in detail by McGeary et. al. (2021) [16]. The final sample size for this trial was N =193.

#### 2.3.1 Inclusion criteria

Completed participation in “Randomized Clinical Trial of Cognitive-Behavior Therapy for Posttraumatic Headache”.

#### 2.3.2 Exclusion criteria

No additional exclusion criteria.

#### 2.3.3 Sample Size and Justification

The sample size for Phase I (model specification) precedes from a fixed, previously collected data set, with N = 129 (n = 65 CBT, n = 64 TAU). This sample size will be sufficient to specify a model using a Random Forest Algorithm, which because of the few number of hyperparameters to tune (see: Biau et al., 2016), has been shown to be performant in similar settings and modest sample sizes (see: Lu et al., 2018).

### 2.4 Outcomes and measures: Phase 1

The outcome of interest was selected to represent the perceived burden of post-traumatic headache. The Headache Impact Test (HIT-6) was selected because it is commonly used [17] and provides a validated estimate of self-reported disability due to headache symptoms. The measure will be scored according to its Item Response Theory (IRT)-based weights, with each item response (Never, Rarely, Sometimes, Very Often, Always) receiving a numerical value (6, 8, 10, 11, 13), resulting in a total score ranging from 36 to 78. The immediate post-treatment score will primarily be used for the development of the prediction model, with all the post-treatment measurements (posttreatment, 3 months, 6 months) also used in a supplemental model. Patient-centered outcome modeling, based on an outcome considered most important by patients with posttraumatic headache, will also be conducted as such information becomes available.

#### 2.4.1. Candidate Predictors

The following baseline (i.e., pre-treatment) predictor candidates were selected based on their theoretical importance and availability in both data sources (see Table 1). This list is subject to additions, if recent literature review or expert opinion suggests that that one or more additional variables present in the database should be included as a candidate predictor.

**Table 1.**
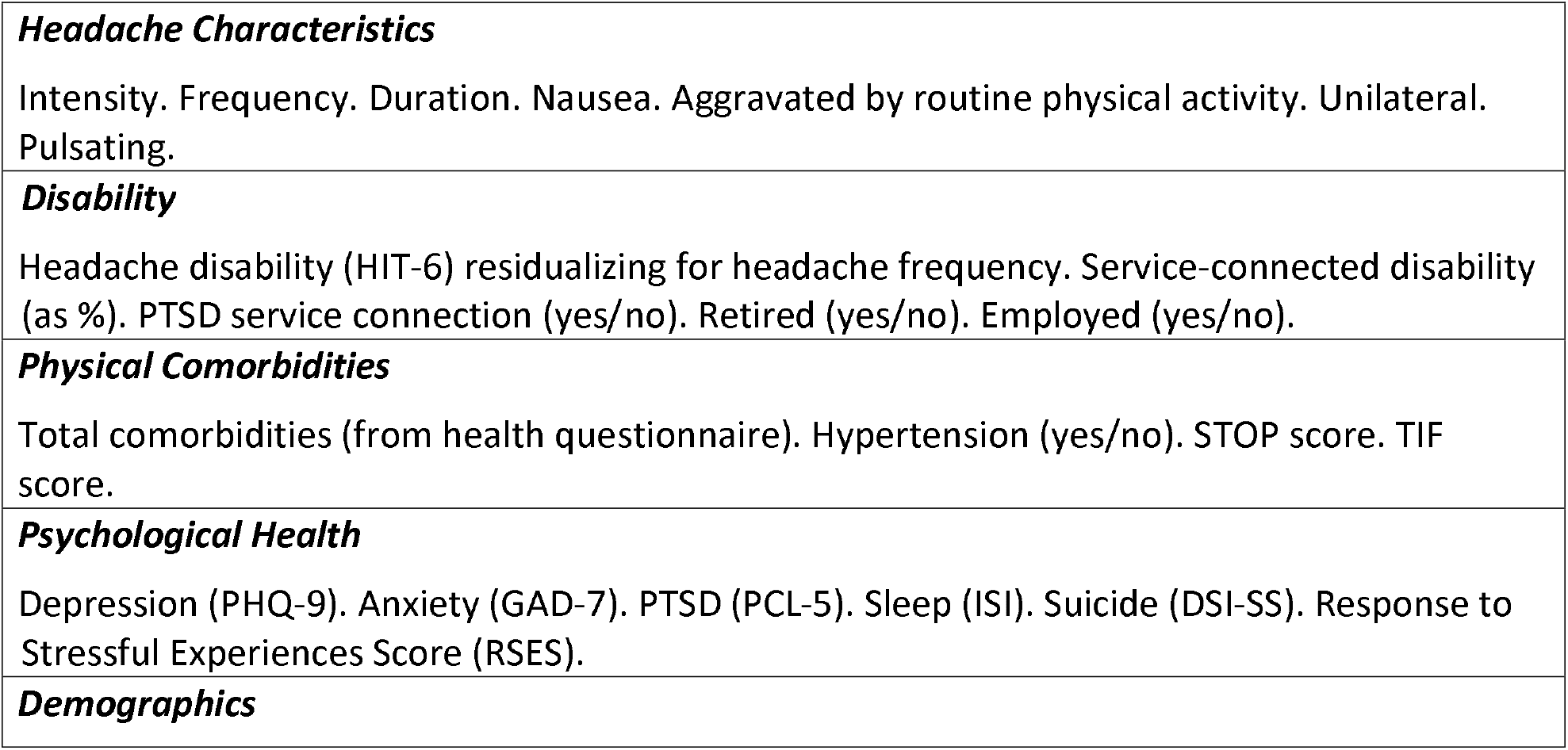
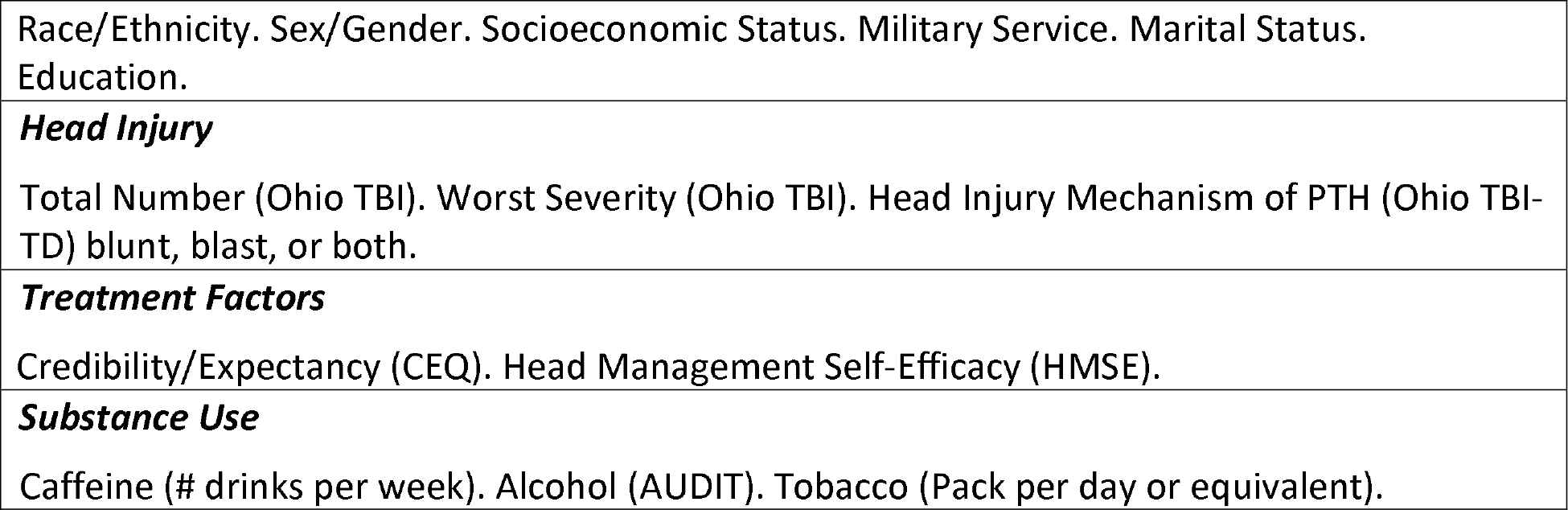
Candidate Predictors.

### 2.5 Model Development: Phase 1

#### 2.5.1 Specification and Estimation

The primary prediction model will be developed with the expectation that pre-treatment patient-level mechanistic and non-mechanistic factors will be associated with future states of patient-centered treatment benefit. The predictor candidates will be evaluated for inclusion in a frequentist generalized linear model with L1-penalized estimation, akin to a LASSO procedure. This estimation procedure shrinks the coefficients of predictors that are not strongly associated with the outcome to zero, effectively removing these variables from the prediction model. For these models, each predictor will be coded using its most informative state (i.e., continuous or ordinal predictors will not be artificially dichotomized for purposes of modeling). Nonlinear relationships using fractional polynomials and interaction terms will also be considered. A Bayesian workflow approach will be used (see Gelman et al., 2020) [18]. We will conduct data simulations in order to better understand the parameters of the model, along with improving our inferences about the priors and determining the probability of observing certain data configurations in future samples.

This approach is described by Kent et al. (2018) as a ‘risk modeling’ approach [19]. In this approach, individuals from both study arms are pooled into a sample for a single model that is blinded to treatment assignment. Modeling the outcome risk in the control arm separately from the treatment arm is alluring, but in the present sample, the risk of overfitting is substantial and may introduce a differential model fit across subsamples based on treatment assignment [19].

#### 2.5.2 Machine Learning Approaches

We will also build several models with supplemental machine-learning approaches. Our intention for these models is twofold: A) We wish to conduct an alternative ‘effect-based’ approach to modeling treatment benefits; B) We wish to explore unimagined patient-level treatment effect benefits using data-driven considerations.

To model the expected treatment effect, we will use what Kent et al. (2018) describe as ‘effect-based’ modeling [19], where the average treatment effect is modeled using observed treatment effects from the derivation trial. For this approach, we will use a nonparametric random forest [20]. This approach has a high probability of overfitting the modest sample, so interpretation will be contingent on the internal cross-validation procedures listed below.

In an additional supplemental model, all the posttreatment outcome assessments will be utilized. For this approach, machine learning methods using a random forest algorithm [21] on our single-site RCT data will also be used to specify the factor-factor and factor-outcome relationships. Additional approaches that are designed to be robust to the clustering of variance expected in individual repeated-measure data patterns for the repeated posttreatment outcome measurements (e.g., maximum likelihood estimation of Gaussian parameters through the ‘rstan’ and ‘tmle’ R packages) will also be applied [22].

#### 2.5.3 Missing Data

For the primary model specification, with one outcome assessment and completely observed predictor variables, there are no missing data in the derivation sample. However, for the supplemental models using all posttreatment assessments, there is missing data for many participants. Further, the external validation data collected through Project MARCH will likely have missing data for some participants.

Where relevant, we will assume the data are missing at random (MAR) conditional on the theoretically selected baseline predictor candidates. Multiple imputation using chained equations (MICE) will be used to impute data missing outcome or predictor data separately for the development and validation data sets. For these procedures m = 50 imputations will be conducted with the estimates combined using Rubin’s rules.

#### 2.5.4 Internal Validation

The derived model(s) will be calibrated and internally validated using bootstrapping or k-fold cross validation [23] in dataset subgroups of patients who completed CBTH and usual care. In the derivation sample, the sample size is quite modest in relation to the number of predictor candidates, so over-optimism will be carefully evaluated, with more parsimonious LASSO solutions given preference over larger models.

#### 2.5.5 Model Evaluation

Several aspects of model performance will be evaluated. Because model calibration is just as important as model discrimination [24], model calibration will be conducted using graphical methods and the evaluation of calibration intercept and slope. Calibration in the large and small will be evaluated based on the total sample and subsets of meaningful subgroups (e.g., sex, age, type of head injury, etc.). Model discrimination for the continuously scaled outcome will be evaluated using variance methods such as R-squared. Additionally, proper score functions will be employed such as the Brier Score. For the supplemental model, the Brier Skill Score is a more appropriate choice for model performance because the Brier Skill Score is more robust to the serial correlation expectation in predictor variables for this Project [25].

## 3.0 Methods: Phase 2 Model Validation

### 3.1 Objective: Phase 2

The model specification and internal and external validation are being conducted on available data from first-of-their-kind RCTs. While a valuable resource, the original RCT was not designed to support a model-building effort like the one proposed. We contend that sufficient information is available to specify and estimate a preliminary model using LASSO procedures but feel less confident that the parameter estimates derived from this sample will be stable and sufficient for future model use. As such, we propose to conduct a hybrid external validation effort using Bayesian methods that will update the parameter estimates based on information from the external sample. In essence, the external model validation serves as both model updating and partial external validation (i.e., a hybrid task).

Once the model is specified, tuned, and internally validated, the analytical team will describe the outcomes, variables, and relationships in the model in plainspoken language for discussion with the investigators. The elements of the model will be discussed, including both predictors and possible measurement issues with these predictors, while seeking input on the selected variables and their measurement. Once complete, the research team will finalize the model specification and prepare the model(s) estimates to be used in the updating and external validation tasks.

The finalized primary model that was estimated using frequentist procedures will be used to prepare prior probability distributions for the external updating and validation model. The point estimates of the mean-centered parameters will serve as the central tendency (i.e., expected value) with 1.5 x the observed standard error for the dispersion parameters to form an informative prior. In this way, the model specification and prior probability distributions can be based on information from the derivation sample. A normal distribution will be assumed as follows:

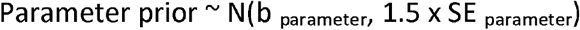

This generalized linear model will be estimated/updated using rstan with priors specified, as above, using the validation sample.

### 3.2 Data Sources: Phase 2

The dataset will be obtained from a second randomized controlled trial (NCT05620719). The data from this study will be stored in a local repository and will be released without identifiers to the study team under a Repository Recipient Investigator Agreement.

### 3.3 Study Population: Phase 2

Model validation will proceed from retrospective data collected from a randomized controlled trial of veterans and active duty military personnel with persistent posttraumatic headache. The eligibility criteria for the analysis are the same as for this individual trial, with no additional exclusion criteria. See clinicaltrials.gov for more information about this trial.

#### 3.3.1 Inclusion criteria

Completed participation in “Multisite Advancement of Research on Chronic Posttraumatic Headache: Project MARCH” (UT Health SA IRB #22-592H).

#### 3.3.2 Exclusion criteria

No additional exclusion criteria.

#### 3.3.3 Sample Size and Justification

For Phase 2 (external validation), a sample size of N = 200 will provide precision of +/-0.10 for R-squared confidence interval (assuming R2 of 0.40) in the calibration slope, so it will allow meaningful evaluation of calibration metrics for the calibration-in-the-large (i.e., comparison of predicted to observed outcome states).

### 3.4 Analytic Approach: Phase 2

In the updating/validation sample, data from N = 200 Project MARCH trial participants (n=100 CBTH and n=100 usual care) will be utilized. The model will be estimated using 4 chains, with 1000 burn-in iterations and 4000 samples per chain. STAN’s No U-Turn sampler will be used, and the chains evaluated for evidence of convergence and unacceptable levels of divergent sampling.

The parameter estimates from the updated model (i.e., priors x likelihood) will be evaluated. Parameter migration (i.e., learning) will be evaluated using Kullback-Leibler divergence with higher levels indicating evidence that the original parameter estimates migrated to new locations in response to the external data (i.e., greater levels of migration are indicative of poor performance using the original parameter space).

Two prognostic indices (i.e., predictions) will be calculated based on the sum of the model predictor variables multiplied by their established predictor weight means (prognostic 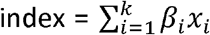. In the first, only the prior probability distributions will be used to estimate the linear predictions. This represents the predicted score based only on the derivation sample information. The second set will use the posterior parameter estimates from the external sample (i.e., the updated model coefficients) to examine prediction. The correspondence of the two predictions will be evaluated using graphical methods to further examine the stability in model prediction across the derivation and updated parameter sets. Model performance, including calibration and discrimination, will be conducted on the updated predictions using the methods described, above.

## 4.0 Concluding Remarks

Treatment of persistent posttraumatic is a challenge that requires advancing and personalizing treatment beyond current practice. The findings of this study are intended to be combined with modeling of pharmacological factors to create a more comprehensive model. This study will significantly contribute to the aim of providing precision medicine for veterans and active-duty military personnel.

## Data Availability

All data utilized for the present study are available up reasonable request to the authors.

## Abbreviations Used

AUDIT: Alcohol Use Disorders Identification Test
CBT: Cognitive-Behavioral Therapy
CEQ: Credibility Expectancy Questionnaire
CSQ-8: Client Satisfaction Questionnaire 8-item
DSI-SS: Depressive Symptom Index – Suicidality Scale
GAD-7: Generalized Anxiety Disorder 7-items
HIT-6: Headache Impact Test 6-items
HMSE: Headache Management Self-Efficacy Scale
IRB: Institutional Review Board
ISI: Insomnia Severity Index
LASSO: Least Absolute Shrinkage and Selection Operator
PATH: Predictive Approaches to Treatment effect Heterogeneity
PCL-5: PTSD Checklist based on DSM-5 criteria
PHQ-9: Patient Health Questionnaire 9-items
PTSD: Posttraumatic Stress Disorder
RCT: Randomized Controlled Trial
RSES: Response to Stressful Experiences Scale
STOP: Sleep measure – Snoring, Tiredness, Observed stop breathing, high blood Pressure
TAU: Treatment as Usual
TBI: Traumatic Brain Injury
TIF: Tinnitus Functional Index Questionnaire

## Additional Contributions

The authors are grateful for the administrative and regulatory support of Deanne Hargita and Anna Gonzalez.

